# Preliminary study of transcranial Doppler observation of cerebral herniation in acute internal carotid artery occlusion

**DOI:** 10.1101/2023.09.05.23295105

**Authors:** Jin Chen, Jun Su, Hong Chang, Peng Bai

## Abstract

**Background and Purpose:** The purpose of this study was to attempt early detection of the risk of cerebral herniation after acute internal carotid artery occlusion (AICAO) using transcranial Doppler.

**Methods:** Twenty-four patients were enrolled within 6 hours of symptom onset. All patients underwent transcranial Doppler (TCD), head computed tomography (CT), magnetic resonance angiography (MRA) or computed tomography angiography (CTA). The National Institutes of Health Stroke Scale (NIHSS) score and the Alberta Stroke Program Early CT Score (ASPECTS) were performed in emergency department and 24 hours after the onset of acute ischemic stroke (AIS) (designated as NIHSS1 and ASPECTS1, respectively). TCD to determine collateral circulation was performed immediately after patient admission.

**Results:** Sex, risk factors, hemisphere, intravenous recombinant tissue plasminogen activator (rt-PA), and time from onset of symptoms to initiation treatment showed no difference between patients with or without cerebral herniation. The age of the group with cerebral hernia was significantly younger than that of the group without cerebral hernia (P=0.000). Assessment of the anterior communicating artery (ACoA) by TCD was in high agreement with assessment by MRA or CTA (kappa=0.625, P=0.002). Correlation analysis showed that the likelihood of a cerebral herniation event was positively correlated with absence of a communicating artery (r_n_=0.660, P=0.002) and negatively correlated with ACoA (r_n_=0.513, P=0.003) and ASPECTS1 (r_s_=0.528, P=0.008), but was not correlated with NIHSS score, NIHSS1 score, △NIHSS (NIHSS1-NIHSS), admission awareness, and stroke volume (P > 0.05).

**Conclusions:** TCD assessment of collateral status could more rapidly indicate the risk of cerebral herniation. Lack of collateral flow may predict the occurrence of cerebral hernia. Compensation of ACoA may indicate a lower risk of cerebral hernia.

## Introduction

Cerebral herniation due to AICAO may result in poor outcomes that can be improved by early decompressive hemicraniectomy (DHC)^1^. Therefore, it is important to identify early factors that may predict cerebral herniation in patients with AICAO. The presence of collateral circulation is thought to be protective against the progression of malignant cerebral edema after ischemic stroke^2^. Therefore, a quick assessment of collateral pathway is essential. Several studies have reported that some imaging techniques such as digital subtraction angiography (DSA), CT angiography (CTA), CT perfusion(CTP), arterial spin labeling(ASL), and MR perfusion(MRP) detect collateral circulation^3,4,5^. In developing or less developed countries, it is difficult to provide some imaging techniques to assess the structure of the cerebral collateral circulation at admission. Therefore, our study focused on safe, noninvasive, and cost-effective bedside test TCD.

The sensitivity and specificity of TCD in detecting the anterior communicating artery were 95% and 100%, respectively, with DSA serving as the reference standard^6^. Several studies^7,8^ have attempted to use TCD or TCCD to assess collateral circulation in AICAO, but no further studies have been performed on the relationship between collateral circulation and cerebral herniation. Our aim was to use TCD to assess collateral circulation in patients with AICAO who progressed to large hemispheric infarction (LHI) to estimate the risk of cerebral herniation.

## Material and methods

This study that included 24 AICAO patients admitted to the neurointensive care unit (NCU) of the Peoples Hospital of Inner Mongolia Autonomous Region between October 2018 and August 2022. The study was approved by the Innermenglia Hospital Ethics Committee (NO.202204212L). All patients underwent standardized treatment according to the AHA/ASA guidelines for the early management of patients with acute ischemic stroke published in 2019^9^.

### The inclusion criteria

1. All patients were admitted within 6 hours of symptom onset and received an acute stroke diagnosis in the emergency department CT.
2. A cerebral focus encompassing at least 2/3 of the ICA territory was confirmed by head CT performed 24 hours after AIS and the volume of the lesions was greater than 100 ml.
3. All patients had no history of cerebral infarction or cerebral infarction that did not result in neurological deficit symptoms or signs.
4. Although several patients received rt-PA intravenous thrombolytic treatment (at a dose of 0.90 mg/kg), the cephalic image score proved ineffective.

### The exclusion criteria

1. Excluded cerebral infarcts with other causes (immune system, infection, blood disease, hereditary vascular disease, patent foramen ovale, dissection, etc.) and cerebral infarcts of unknown cause.
2. Poor temporal window penetration, unable to assess collateral circulation by TCD.
3. Excluded neuroimaging in hemorrhagic transformation.

### Endpoint

Neurologic deterioration consisting of an increase in NIHSS score of > 2 points and a decrease in level of consciousness and a skull scan showing a midline shift of > 5 mm at the septum pellucidum or pineal gland, which is defined as brain herniation. The patients were divided into cerebral hernia group and non-cerebral hernia group according to whether or not they had cerebral hernia.

### Clinical evaluation and data collection

Baseline data, including demographic characteristics and risk factors, were collected immediately after patient admission. The NIHSS score and ASPECTS were performed in the emergency department and 24 hours after the onset of AIS (labeled NIHSS1 and ASPECTS1).

### Head imaging examination

A head examination CT was performed in the emergency department and the second head examination CT was performed 24 hours after AIS onset. The second CT was used to assess ASPECTS1 (ASPECTS1 was independently assessed by two experienced neurology physicians) and calculate infarct volume. After the patient was admitted to the NCU ward, magnetic resonance imaging (MRI), MRA examination, or CTA were performed as soon as possible.

### Assessment of collateral circulation

Detected collateral channels through ACoA, posterior communicating arteries (PCoA), and the external carotid artery (ECA) to ICA collateral circulation compensation levels were evaluated by TCD immediately after patient admission. The use of TCD according to the Chinese Guidelines for Vascular Ultrasound for Stroke published in 2015^10^is to detect collateral channels.

### Collateral flow through the ACoA

1. A decrease in mean flow velocity (MFV) and pulsatility in the ipsilateral middle cerebral artery (MCA) along with normal flow in the contralateral MCA.
2. No significant change in MFV of the ipsilateral MCA and A1 ACA by compression test of ipsilateral common carotid artery, but decreased MFV by compression test of contralateral common carotid artery (CCA).
3. Increased blood flow in the contralateral A1 ACA.

### Collateral flow through the PCoA

An apparent asymmetry of blood velocity of the PCAs from side to side with a high blood velocity ipsilateral to the ICA occlusion.

### Collateral flow from the ECA to the ICA

The supratrochlear artery is a branch of the OA. It can be detected with a 4Hz probe at the medial canthus. The MFV of the supratrochlear artery (STA) was significantly reduced by compression of the ipsilateral superficial temporal artery and mandibular artery, which are branches of the ECA, indicating the formation of collateral channels from the ECA to the ICA.

## Statistics

The distribution of the data was visualized using the Kolmogorov-Smirnov test. Data with normal distribution were presented as mean, range, and SD, whereas data with nonnormal distribution were presented as median, range, and interquartile range. All categorical data were presented as percentages or absolute numbers. Categorical variables were analyzed with Fishers test, student’s t-test, and Mann Whitney U test, as appropriate. Consistency of collateral circulation assessment was analyzed using kappa consistency analysis. Correlation analysis with the Spearman or Pearson correlation coefficient was used to determine the univariate associations between variables. Statistical analysis was performed with SPSS 22.0 software (IBM Corp, Armonk, NY, USA). Results were considered significant at P<0.05.

## Results

### Basic information about AICAO patients

Twenty-four patients were studied (mean age, 69.0±1 2.5years), median baseline NIHSS score was 15.7 (interquartile range, 10–22), median NIHSS1 score and ASPECTS1 24hours after onset were 17.7 (interquartile range, 13–23) and 3.3 (interquartile range, 1–6), and the median infarct volume was 228.08ml (interquartile range, 100–396ml).Regarding blood flow, sixteen patients (67%) had collateral blood flow, eight patients (33%) had ACoA, six patients (25%) had PCoA only, and two patient (8.3%) had collateral blood flow from ECA to ICA only(Table 1).

**Table 1.** Demographic and Clinical Characteristics of the Study Population.

| Characteristic |  |
| --- | --- |
| NO. | 24 |
| Age, y, mean (range, SD) | 69.0 (47–88, 12.5) |
| Sex, males, no. (%) | 18 (75) |
| Smoking history, no. (%) | 10 (41.7) |
| Atrial fibrillation, no. (%) | 4 (16.7) |
| Diabetes, no. (%) | 10 (41.7) |
| Hypertension, no. (%) | 20 (83.3) |
| Stroke volume, ml, median (range, IQR) | 228.08 (100–396, 99.9) |
| NIHSS at admission,median (range,IQR) | 15.7(10-22,3.6) |
| NIHSS1 24h after AIS onset,median (range,IQR) | 17.7(13-23,2.9) |
| ASPECTS1 24h after AIS onset,median (range,IQR) | 3.3(1-6,1.5) |
| cerebral hernia,no.(%) | 10(42) |
| ACoA,no.(%) | 2(8.3) |
| ACoA+PCoA,no.(%) | 4(17) |
| ACoA+ECA to ICA,no.(%) | 2(8.3) |
| ECA to ICA,no(%) | 2(8.3) |
| PCoA,no.(%) | 6(25) |
| Intravenous rtPA,no.(%) | 18(75) |

There were no differences between the group with cerebral hernia and the group without cerebral hernia in terms of sex, risk factors, hemisphere, intravenous rtPA, time from onset of symptoms to initiation treatment. The age of the group with cerebral hernia was significantly younger than that of the group without cerebral hernia (P=0.000) (Table 2).

**Table 2.** Characteristics of AICAO Patients with and without Cerebral Hernia.

| Parameters | Cerebral<br>hernia(n=10) | No cerebral<br>hernia(n=14) | P |
| --- | --- | --- | --- |
| **Demographics** |  |  |  |
| Age | 59.60±9.53 | 75.71±9.42 | 0.000 |
| Sex (male %) | 6(60.0%) | 12(85.7%) | 0.192 |
| **Risk factors** |  |  |  |
| Smoking history (%) | 4(40.0%) | 6(42.9%) | 1.000 |
| Atrial fibrillation (%) | 2(20.0%) | 4(28.6%) | 1.000 |
| Diabetes (%) | 4(40.0%) | 6(42.9%) | 1.000 |
| Hypertension (%) | 8(80.0%) | 12(85.7%) | 1.000 |
| **Clinical features** |  |  |  |
| Hemisphere(right) | 8(80.0%) | 10(71.4%) | 1.000 |
| NIHSS score at admission | 15.40±3.75 | 15.86±3.48 | 0.762 |
| NIHSS score 24h after onset(NIHSS1) | 18.20±3.61 | 17.29±2.33 | 0.459 |
| ΔNIHSS score(NIHSS1-NIHSS) | 2.80±3.22 | 1.43±2.21 | 0.263 |
| ASPECTS 24h after onset(ASPECTS1) | 2.40±1.07 | 4.00±1.47 | 0.008 |
| ΔASPECTS score(ASPECTS1-ASPECTS) | 7.6±1.07 | 6.0±1.47 | 0.008 |
| Stroke volume | 260.6±84.76 | 204±102.69 | 0.174 |
| Level of consciousness at admission | 6.1 | 6.79 | 0.723 |
| Time from symptom onset to treatment start(min) | 108.80±72.17 | 93.43±64.18 | 0.588 |
| **Treatment** |  |  |  |
| Intravenous rtPA | 8(80.0%) | 10(71.4%) | 1.000 |

### The number of collaterals in the groups with cerebral hernia and without cerebral hernia

None of the eight patients with ACoA had a cerebral hernia. Six patients had PCoA only, four patients had no cerebral hernia and two patients had cerebral hernia. Two patients with ECA to ICA collateral circulation had no cerebral hernia. All eight patients had cerebral hernia without collateral circulation. (Figure)

**Figure.**
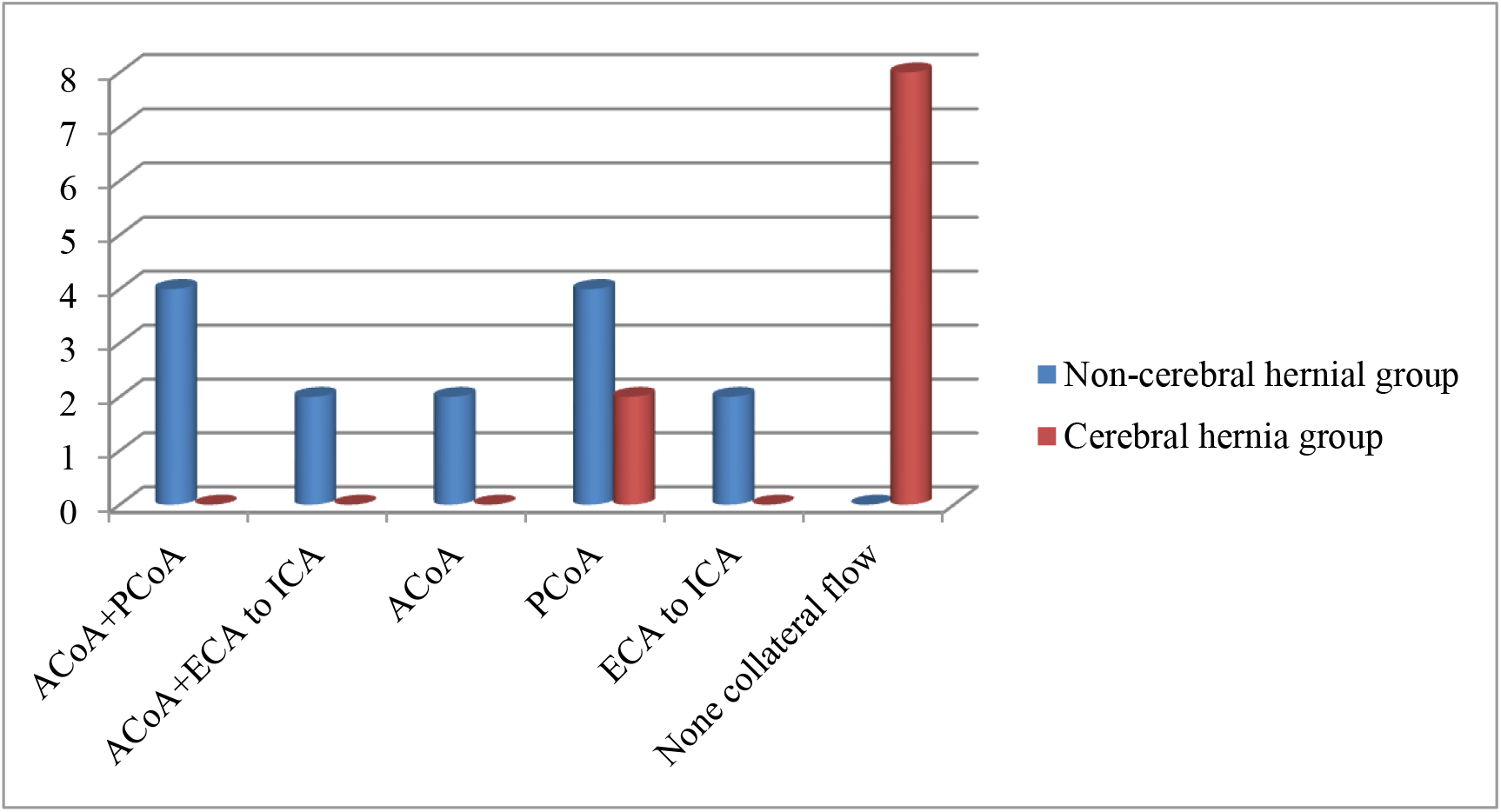
The number of cases of collateral in cerebral hernia and non-cerebral hernia groups.

### Consistency analysis for ACoA assessment and correlation analysis of cerebral hernia events

The assessment of ACoA by TCD was highly consistent with the assessment by MRA or CTA (kappa=0.625, P=0.002) (Table 3). Correlation analysis showed that the probability of cerebral herniation was positively related to the absence of a communicating artery (r_n_=0.660, P=0.002) and negatively related to the anterior circulation balance (r_n_=0.513, P=0.003) and ASPECTS1 (r_s_=0.528, P=0.008) (Table 4) but was not associated with NIHSS score, NIHSS1 score, △NIHSS, admission awareness and stroke volume (P > 0.05).

**Table 3.** The consistency of ACoA measured by CTA and TCD.

| Collateral Flow | ACoA | Kappa | P |
| --- | --- | --- | --- |
| Angiography and TCD showed opened ACoA | 6 | 0.625 | 0.002 |
| TCD showed opened ACoA | 2 |  |  |
| Angiography showed opened ACoA | 2 |  |  |
| Angiography and TCD no showed opened ACoA | 14 |  |  |

**Table 4.** Correlations between Collateral Flow/ASPECTS1 and Cerebral hernia.

| **Collateral Flow/ ASPECTS1** | **$r_n / r_s$** | **P** |
| --- | --- | --- |
| No collateral circulations | 0.660 | 0.002 |
| ACoA | -0.513 | 0.003 |
| ASPECTS1 | -0.528 | 0.008 |

## Discussion

Collateral flow from ACoA, PCoA, ECA to MCA can be assessed with TCD. This is because it can precisely measure the velocity of cerebral blood flow^11^. In this study, we used this technique to assess the status of collateral blood flow after AICAO. The results of the study showed that TCD was in assessing the emergence of collateral blood flow after AICAO. In addition, this procedure is convenient and very inexpensive compared with other methods.

In this study, the systolic flow velocity of the middle cerebral artery on the affected side was 31.45-100.02cm/s when the anterior communicating branch was open, and the systolic flow velocity of the middle cerebral artery on the affected side was only 24% on the opposite side in one of the patients, this patient did not develop malignant cerebral edema. Thus, we believe that the risk of cerebral herniation is very low as long as the ACoA is open. All patients without collateral circulation had cerebral hernia. Because of the small number of cases, regression models could not be constructed to evaluate the risk of cerebral hernia without collateral circulation. We will collect more cases in the future. There were six patients in the posterior communication branch, of which two had cerebral hernia and four did not have cerebral hernia. When compared with the head CT of these six patients, it was found that the width of the central sulci and superior frontal sulci in the normal brain tissue of the four patients without cerebral hernia was wider than that of the two patients with cerebral hernia. In two patients with collateral circulation from the ECA to the ICA, no cerebral hernia formed, and the head CT of these patients showed widening of the sulci in the occipital lobe and marked brain atrophy. This seems to be consistent with the findings of this study that the age of patients with cerebral hernia is younger than that of patients without cerebral hernia.

In this study, the ASPECTS1 was likely to predict cerebral herniation 24 hours after AIS onset. Although it was found that NIHSS score, NIHSS1 score, △NIHSS, ASPECTS, and consciousness did not play a role in the risk assessment of cerebral hernia, this may be related to the fact that the symptoms of neurological impairment had not yet progressed to the point that patients sought medical attention early (median 99min).This is consistent with the findings of Takashi Shimoyama et al^12,13^, but contradicts the results of certain clinical trials in which patients with acute cerebral infarction were evaluated within 24 hours of symptom onset^14,15^.

It is expected that TCD will allow for the first time to assess the establishment of the collateral circulation in patients with massive cerebral infarction due to acute occlusion of the internal carotid artery, which may allow prediction of the occurrence of cerebral herniation and provide a basis for predicting the direction of development of cerebral edema.

## Data Availability

All data analyzed during this study are included in this article.

